# Understanding spatiotemporal clustering of seasonal influenza in the United States

**DOI:** 10.1101/2025.05.28.25328505

**Authors:** Louis Yat Hin Chan, Sinead Morris, Norman Hassell, Perrine Marcenac, Alexia Couture, Arielle Colon, Krista Kniss, Alicia Budd, Matthew Biggerstaff, Rebecca Borchering

**Author notes:** Corresponding author; email: Influenza Division, Centers for Disease Control and Prevention, 1600 Clifton Road, Atlanta, GA 30329, USA.

## Abstract

**Background:** Seasonal influenza exhibits distinct spatiotemporal patterns across the United States, yet the geographic clustering of influenza activity remains incompletely understood. This study aims to identify jurisdictions with similar patterns of seasonal influenza epidemics by exploring spatiotemporal dynamics across the United States after the 2009 H1N1 pandemic.

**Methods:** We analyzed data from U.S. influenza surveillance systems, including outpatient illness surveillance and virologic surveillance. The outpatient illness data included weekly proportions of outpatient visits for influenza-like illness from jurisdictions including all 50 states, while virologic data comprised influenza test positivity results from U.S. public health and clinical laboratories covering all 50 states. We calculated Moran’s I statistics to assess spatial autocorrelation in peak timing. We also performed k-means clustering on z-normalized time series data and determined optimal clusters using the silhouette method. We then conducted an analysis of variance (ANOVA) to evaluate differences among clusters based on the Moran’s I statistics and the relative proportions of influenza virus types and subtypes.

**Results:** Our analysis revealed distinct spatial clusters with significant geographic patterns. We found a consistent grouping of Southeastern states (Georgia, Alabama, Mississippi, Louisiana, and Florida). This clustering pattern was partially explained by earlier seasonal peaks in these jurisdictions and supported by significant spatial autocorrelation in peak timing. While Southeastern states maintained stable cluster associations, Western and Central states showed greater variation in cluster membership across seasons. We also found significant differences between clusters in the Moran’s I statistics and the proportion of all influenza A virus detections that were influenza A/H1 viruses. However, no significant differences were found in the proportion of all influenza A and B virus detections that were influenza A viruses.

**Conclusions:** These findings quantify the distinct spatiotemporal patterns of seasonal influenza in the Southeastern United States compared to other regions. Understanding these regional clustering patterns can enhance preparations for upcoming changes in influenza activity and inform targeted public health interventions such as timing of vaccination campaigns. Robust surveillance systems, adaptive approaches, and stable long-term data are essential for effectively addressing regional differences and ultimately strengthening nationwide preparedness for seasonal influenza.

## Introduction

Seasonal influenza remains a major public health concern worldwide, characterized by substantial morbidity, mortality, and economic impacts (Iuliano, et al. 2018) (Putri, et al. 2018) (Hayward, et al. 2014) (de Courville, et al. 2022) (Couture, et al. 2025). Influenza viruses are primarily classified into influenza types A and B, with influenza A viruses (subtypes A/H1 and A/H3) generally causing the majority of infections (Sumner, et al. 2023). The burden of seasonal influenza in the United States is significant and varies by season (Centers for Disease Control and Prevention 2025). For example, the 2017/2018 influenza season alone resulted in an estimated 35–52 million symptomatic illnesses, 550,000–1 million hospitalizations, and 36,000–98,000 deaths (Centers for Disease Control and Prevention 2025).

Influenza outbreaks exhibit marked temporal patterns, with activity typically beginning to increase in November and peaking between December and February in the United States (Centers for Disease Control and Prevention 2025). While these temporal dynamics have been well-studied (Nsoesie, et al. 2014), revealing consistent seasonality across years, there remains a significant research gap in whether seasonal influenza epidemics vary by geography within the United States. Surveillance data indicate that seasonal influenza epidemics in the United States frequently, but not always, originate in the Southeastern region, particularly in states such as Florida and Georgia, before spreading to other areas. However, these spatial patterns are dynamic and vary annually due to complex interactions between climate, population density, human mobility, and viral evolution (Shaman and Kohn 2009) (Shaman, Pitzer, et al. 2010) (Dalziel, et al. 2018) (Chen, et al. 2024) (Viboud, et al. 2006) (Charu, et al. 2017). This variability complicates efforts to predict where influenza activity will increase next and presents challenges for effective preparedness and response to seasonal influenza outbreaks.

Several studies have contributed to our understanding of influenza spatial dynamics in the United States. Gog et al. (Gog, et al. 2014) and Kissler et al. (Kissler, et al. 2019) demonstrated that the fall wave of the 2009 influenza pandemic activity originated in the Southeastern United States, while Charu et al. (Charu, et al. 2017) identified clear spatial patterns in influenza onset times across the United States from 2002 to 2010, with most seasons starting in the Southern United States. Rosensteel et al. (Rosensteel, et al. 2021) further highlighted significant heterogeneity in regional clustering patterns across the seasons from 2002 to 2009. In contrast, Dahlgren et al. (Dahlgren, et al. 2019) found no evidence of meaningful spatial autocorrelation in peak timing for the 2010–2016 seasons. Despite these insights, there remains limited understanding of how spatial patterns have evolved over time, particularly in the years following the 2009 H1N1 pandemic.

Spatial clustering refers to the grouping of regions with similar patterns of influenza activity, which can provide insights into the spread of influenza within a country. By examining these spatiotemporal dynamics, we can enhance our understanding of influenza spread and the variability in regional activity. This study examines the spatial clustering of influenza activity across the United States after the 2009 H1N1 pandemic. We first analyze spatiotemporal dynamics by examining the peak timing of time series data from influenza outpatient illness and virologic surveillance systems (Centers for Disease Control and Prevention 2024). Then, we identify spatial clusters by applying k-means clustering, an unsupervised machine learning technique, to the time series data. Finally, we apply analysis of variance (ANOVA) to explain the regional differences in influenza dynamics. Our findings contribute to the broader field of spatiotemporal epidemiology and offer insights that can help inform prevention and control efforts for future influenza seasons.

## Methods

### Surveillance data and preprocessing

This study utilized two primary surveillance systems: outpatient illness surveillance and virologic surveillance. First, we used unweighted weekly proportions of outpatient visits for influenza-like illness (ILI), defined as fever with cough or sore throat, from the U.S. Outpatient Influenza-like Illness Surveillance Network (ILINet). These data may capture a range of respiratory infections caused by pathogens other than influenza viruses. Second, we used weekly values of the percentage of specimens testing positive for influenza virus from virologic surveillance, which includes data from the U.S. World Health Organization (WHO) Collaborating Laboratories System and the National Respiratory and Enteric Virus Surveillance System (NREVSS). These data included test results from approximately 100 public health laboratories, which test respiratory specimens for surveillance purposes, and 300 clinical laboratories, which focus on diagnostic testing. For the period before the 2015/2016 season, both public health and clinical laboratory data were reported together. However, from the 2015/2016 season onward, they were reported separately (Centers for Disease Control and Prevention 2024), with weekly values at the state and territory level available only for the clinical laboratory data.

All data from the 2010/2011 to 2023/2024 seasons were sourced from FluView Interactive (Centers for Disease Control and Prevention 2024), with the weekly data collected by epidemiologic week, referred to as MMWR Week (Centers for Disease Control and Prevention 2025). In this study, each season was defined as beginning in Week 40 (around early October) each year and ending in Week 39 the following year. The study period focused on two phases: the pre-COVID-19 period (2010/2011–2019/2020) and the post-COVID-19 period (2022/2023–2023/2024). All 50 states, Puerto Rico, and the District of Columbia were covered in both surveillance systems. The outpatient illness surveillance also included the U.S. Virgin Islands and New York City, for a total of 54 jurisdictions. New York City and New York state were reported as separate jurisdictions and treated as distinct entities for the purposes of this study. As a result, the U.S. Virgin Islands and New York City were excluded from analyses using the virologic surveillance data. Data from the territories of American Samoa, Guam, and the Northern Mariana Islands were not available in either surveillance system.

Missing weekly values (the unweighted weekly proportions of outpatient visits for ILI and the weekly percentages of specimens testing positive for influenza) occurred in both datasets, with the latter particularly affected during periods outside the core influenza season due to lower testing volume and variability in voluntary reporting across jurisdictions (Figure S1). For example, Idaho reported the weekly percentages of specimens testing positive for influenza only during the core influenza period, and New Jersey reported these data only for the 2023/2024 season. We imputed the missing weekly values for each jurisdiction using Stineman interpolation, implemented with the ‘imputeTS’ package in R (Moritz, et al. 2022). Jurisdictions with more than 50% missing weekly values within a season were excluded from that season in our analyses.

To reduce noise and elucidate the temporal patterns in the data, we applied smoothing using Nadaraya– Watson kernel regression (Brockwell and Davis 1991). This step ensured that seasonal trends were more discernible by minimizing short-term fluctuations. To analyze trends over a period of *T* seasons, we then averaged the smoothed data across seasons for each jurisdiction as follows

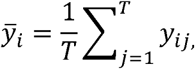

where *ȳ_i_* is the average value for week *i*, and *y_ij_* is the value for week *i* in season *j*. We note that data for week 53 were excluded from the averaged dataset but were kept when analyzing individual seasons. The weekly data (for individual seasons and averaged across seasons) were used in subsequent peak timing spatial analysis and time series clustering analysis.

Finally, the public health data from the virologic surveillance system provided raw detection counts for each influenza virus type (A and B) and subtype (A/H1 and A/H3) by season and jurisdiction. We calculated the seasonal proportions of each type and subtype and included these as dependent variables when exploring cluster differences in a subsequent ANOVA.

### Peak timing spatial analysis

We calculated global and local Moran’s I statistics for each dataset (the unweighted weekly proportions of outpatient visits for ILI and the weekly percentages of specimens testing positive for influenza) to investigate the spatial autocorrelation in timing of peak influenza activity. Moran’s I statistics evaluate the degree to which the values in each jurisdiction are similar to those in neighboring jurisdictions, providing a measure of spatial clustering. For this analysis, only continental U.S. jurisdictions (including New York City) were included, while jurisdictions such as Alaska, Hawaii, Puerto Rico, and the U.S. Virgin Islands were excluded.

To measure spatial autocorrelation across the United States, we calculated the global Moran’s I statistic and its associated p-value using a permutation test implemented through the ‘spdep’ package in R (Bivand, et al. 2025). Positive values indicate positive spatial autocorrelation or clustering, where jurisdictions with similar peak timing are geographically close to one another. Values near zero suggest a random spatial distribution with no discernible pattern, while negative values indicate negative spatial autocorrelation or dispersion, where jurisdictions with dissimilar peak timing are more likely to be neighbors. The formula for the global Moran’s I is

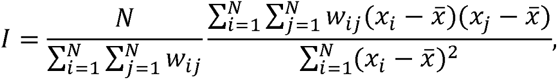

where *N* is the total number of jurisdictions, *w_ij_* represents the spatial weight between jurisdictions *i* and *j, x_i_* is the peak timing at jurisdiction *i* and *x* is the average peak timing across all jurisdictions.

In addition to the global Moran’s I, we calculated local Moran’s I for each jurisdiction to identify localized patterns of spatial autocorrelation, such as differences within specific clusters or the presence of outliers. The local Moran’s I was also computed using the ‘spdep’ package in R (Bivand, et al. 2025). The formula for the local Moran’s I is

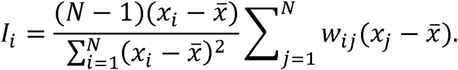

### Time series clustering analysis

We performed a k-means clustering analysis using the ‘dtwclust’ package in R (Sarda-Espinosa 2024). The analysis aimed to group jurisdictions with similar influenza activity patterns over time, ensuring that time series within the same cluster followed comparable temporal trends while remaining distinct from those in other clusters. To quantify differences in temporal trends, we used the Manhattan L1 distance measure.

To ensure comparability across jurisdictions and seasons, we first applied z-normalization to standardize the smoothed time series data and place all data on a consistent scale. For k-means clustering, we then used the silhouette method (Rousseeuw 1987) to determine the number of clusters (*k*) that maximizes the separation between clusters while minimizing the variance within clusters. We explored a range of potential cluster counts up to a maximum of *k* = 10. We calculated the silhouette score for each value of *k* and selected the *k* with the highest silhouette score.

To ensure robustness, we repeated the clustering 100 times with different random initializations (i.e., different random seeds) and selected the solution with the highest consistency across these runs. Results of single-jurisdiction clusters were manually excluded, as they tended to overfit the data and did not represent meaningful spatial groupings. We also examined the dispersion of jurisdictions within each cluster, calculating the average intra-cluster Manhattan L1 distance in terms of standard deviations, to identify outliers and assess the clustering quality. This metric helped assess whether jurisdictions within the same cluster were closely grouped or exhibited substantial variability.

We conducted the above clustering analysis by first stratifying each dataset (the unweighted weekly proportions of outpatient visits for ILI and the weekly percentages of specimens testing positive for influenza) separately in univariate analyses. Then, we assessed joint clustering patterns by performing a multivariate analysis that combined both datasets. Note that in the multivariate analysis, a jurisdiction had to be available in both datasets in order to be included (e.g., New York City was excluded because the weekly percentages of specimens testing positive for influenza were not available).

### Evaluating cluster differences using ANOVA

Following the peak timing spatial analysis and time series clustering analysis, we conducted an ANOVA to assess differences among clusters based on four dependent variables: peak timing, local Moran’s I, the proportion of all influenza A and B virus detections that were influenza A viruses, and the proportion of all influenza A virus detections that were influenza A/H1 viruses. This analysis was performed on clustering patterns across individual seasons (not averaged across seasons) to capture season-to-season variations, and we focused on the pre-COVID-19 era (2010/2011–2019/2020) to exclude differences associated with pandemic-related disruptions. The model included three effects: cluster, season, and dataset, with the interaction between these effects also considered. The model was specified as

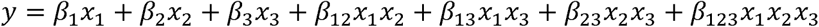

where *y* is the dependent variable, *x*_·_ represent the effects, and *β* represent the model coefficients. To account for multiple comparisons (McHugh 2011), we then performed a post hoc analysis using the Tukey’s honestly significant difference (Tukey HSD) test (Yandell 2017). Further details are provided in the supplementary materials.

## Results

### Preprocessing surveillance data

We included data from all 54 jurisdictions for most seasons in the unweighted weekly proportions of outpatient visits for ILI from the outpatient illness surveillance, with two exceptions: data from the U.S. Virgin Islands (VI) were excluded for the 2010/2011 season, and data from Puerto Rico (PR) were excluded for three seasons (2010/2011–2012/2013). For the weekly percentages of specimens testing positive for influenza from the virologic surveillance, fewer jurisdictions were included (ranging from 40 to 47 out of 54), as several jurisdictions had more than 50% missing weekly values in some seasons (Figure S1). All seasons were excluded for the District of Columbia (DC), New Jersey (NJ) and Rhode Island (RI) due to insufficient data. Approximately 6% of the weekly values were imputed in the included jurisdictions (Table S1).

### The Southeastern states experienced earlier peak timing

Our analysis revealed that peak timing, based on the averaged time series data for the pre-COVID-19 period (2010/2011–2019/2020), occurred earlier in the Southeastern states for both datasets (the unweighted weekly proportions of outpatient visits for ILI and the weekly percentages of specimens testing positive for influenza) (Figure 1). Specifically, Georgia (GA) and Alabama (AL) peaked approximately 5 weeks earlier than the averaged peak timing across all jurisdictions. Additionally, some Western states also exhibited earlier peak timing. For example, Utah (UT) peaked approximately 4 weeks earlier. In contrast, Montana (MT) experienced peak timing about 4 to 5 weeks later than the average. While both datasets demonstrated similar spatial patterns, slight variations were observed between them. For example, the weekly percentages of specimens testing positive for influenza virus showed less consistency in peak timing compared to the unweighted weekly proportions of outpatient visits for ILI, with a broader range of peak weeks across states, particularly in the Central and Northeastern states. For the post-COVID-19 period (2022/2023–2023/2024), the peaks generally occurred earlier than in the pre-COVID-19 period.

**Figure 1.**
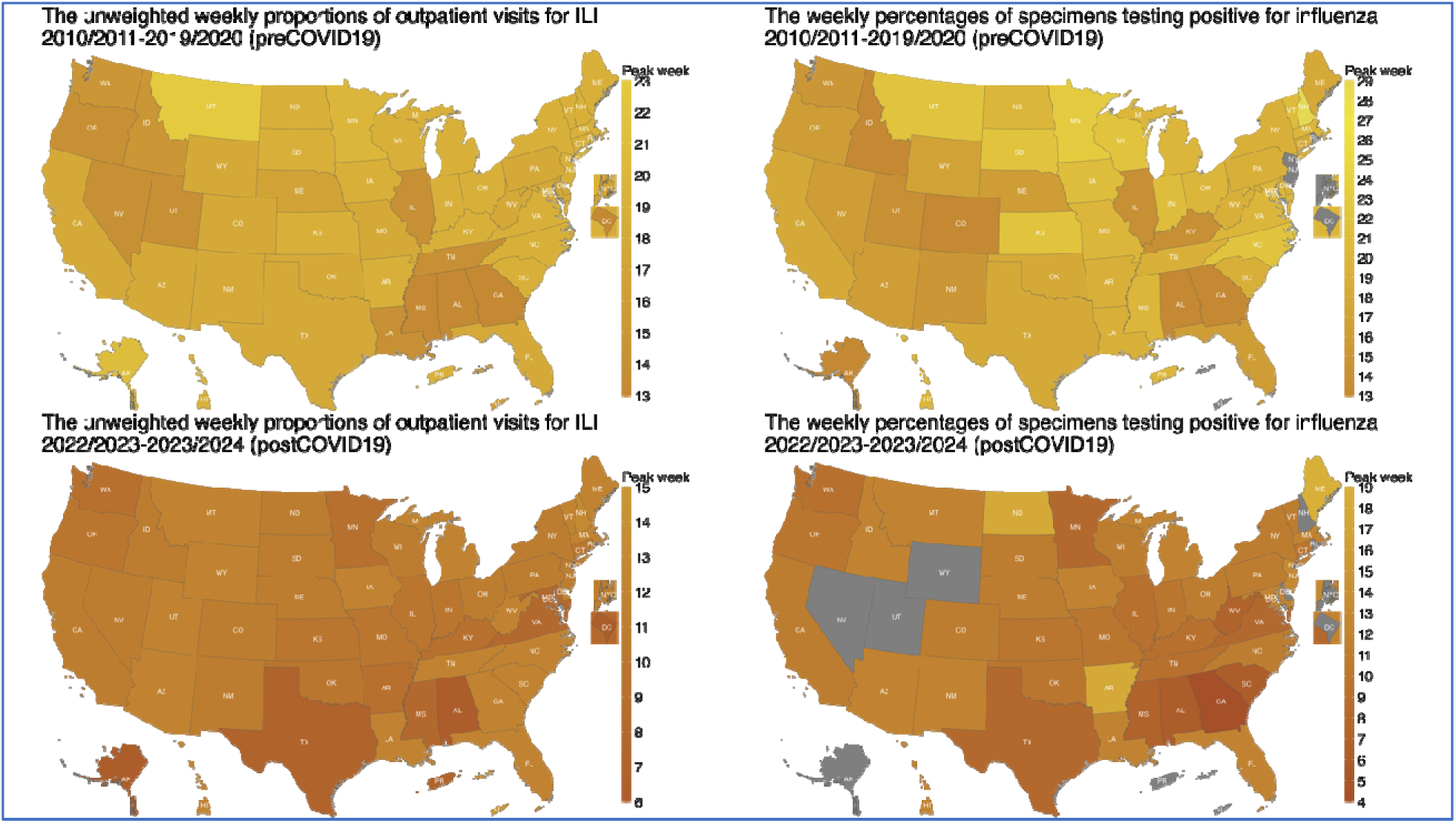
Spatial distribution of peak timing for the unweighted weekly proportions of outpatient visits for ILI (left panels) and the weekly percentages of specimens testing positive for influenza (right panels), averaged across seasons. The latter consist of both public health and clinical laboratory data reported before the 2015/2016 season, while only the clinical laboratory data were available from the 2015/2016 season onward. The upper panels represent the pre-COVID-19 seasons (2010/2011– 2019/2020), while the lower panels represent the post-COVID-19 seasons (2022/2023–2023/2024). Darker yellow indicates earlier peak timing, while lighter yellow represents later peak timing. All panels share the same color scale, which corresponds to the week from the beginning of influenza season (MMWR Week 40). The gray color (e.g., in DC, NJ, and NYC) indicates that all seasons were excluded from the analysis due to missing data.

For the pre-COVID-19 period, the global Moran’s I values were 0.29 for the unweighted weekly proportions of outpatient visits for ILI and 0.30 for the weekly percentages of specimens testing positive for influenza, with both p-values < 0.05, indicating statistically significant spatial autocorrelation (Table 1). These results suggest that jurisdictions with similar peak timings tend to cluster geographically.

**Table 1.** Global Moran’s I values (with p-values in parentheses) of peak timing for the unweighted weekly proportions of outpatient visits for ILI and the weekly percentages of specimens testing positive for influenza, across multiple seasons and single seasons. The latter consist of both public health and clinical laboratory data reported before the 2015/2016 season, while only the clinical laboratory data were available from the 2015/2016 season onward. Bold values indicate statistically significant spatial autocorrelation (p < 0.05).

| Study period | The unweighted weekly proportions of outpatient visits for ILI | The weekly percentages of specimens testing positive for influenza |
| --- | --- | --- |
| Multiple seasons |  |  |
| 2010/2011–2019/2020 | **0.29 (<0.05)** | **0.30 (<0.05)** |
| 2022/2023–2023/2024 | **0.20 (<0.05)** | 0.04 (0.25) |
| **Single season** |  |  |
| 2010/2011 | **0.27 (<0.05)** | **0.38 (<0.05)** |
| 2011/2012 | 0.00 (0.39) | 0.08 (0.13) |
| 2012/2013 | **0.45 (<0.05)** | -0.03 (0.35) |
| 2013/2014 | **0.36 (<0.05)** | **0.13 (<0.05)** |
| 2014/2015 | **0.60 (<0.05)** | -0.04 (0.56) |
| 2015/2016 | 0.08 (0.15) | 0.11 (0.11) |
| 2016/2017 | **0.49 (<0.05)** | **0.17 (<0.05)** |
| 2017/2018 | **0.42 (<0.05)** | **0.25 (<0.05)** |
| 2018/2019 | **0.18 (<0.05)** | **0.28 (<0.05)** |
| 2019/2020 | -0.01 (0.43) | **0.19 (<0.05)** |
| 2020/2021 | -0.08 (0.71) | 0.02 (0.31) |
| 2021/2022 | 0.05 (0.19) | **0.25 (<0.05)** |
| 2022/2023 | **0.60 (<0.05)** | **0.34 (<0.05)** |
| 2023/2024 | **0.20 (<0.05)** | **0.21 (<0.05)** |

Further analysis using the local Moran’s I revealed distinct localized spatial patterns between the two datasets (Figure S2). For the unweighted weekly proportions of outpatient visits for ILI, higher local Moran’s I values were observed in Southeastern states such as Alabama (AL), while for the weekly percentages of specimens testing positive for influenza, higher values were found in Western states such as Utah (UT). These findings highlight specific regions where clustering of peak timing is stronger and demonstrate that spatial associations varied across jurisdictions and datasets. In contrast, our analysis for the post-COVID-19 period revealed distinct differences compared to the pre-COVID-19 period. The global Moran’s I values were lower for both datasets, indicating weaker spatial autocorrelation (Table 1).

We also examined peak timing during individual seasons (Figure S3 and Figure S4). We found that the global Moran’s I values varied across seasons (Table 1), with more than half showing statistically significant spatial autocorrelation (p-values < 0.05). This suggests that while spatial clustering of peak timing is a consistent feature overall, its strength fluctuates from season to season. In particular, the global Moran’s I values were not significant for the 2020/2021 season, due to notably lower influenza activity during the COVID-19 pandemic. However, the 2022/2023 and 2023/2024 seasons showed higher global Moran’s I values with p-values < 0.05, indicating the re-emergence of spatial clustering and a gradual return to more typical spatial patterns of influenza activity. Similarly, the local Moran’s I values also exhibited seasonal variability, reflecting changes in the strength and patterns of localized spatial clustering (Figure S5 and Figure S6).

### Consistent clustering patterns in the Southeastern states

The time series clustering analysis identified two distinct spatial clusters, indicating that influenza activity across the United States primarily followed two comparable temporal trends (Figure 2). One of the clusters was a Southeastern cluster, which consistently included 5 core members: Georgia (GA), Alabama (AL), Mississippi (MS), Louisiana (LA), and Florida (FL). During the pre-COVID-19 period (2010/2011–2019/2020), the Southeastern cluster also included Texas (TX) and Puerto Rico (PR).

**Figure 2.**
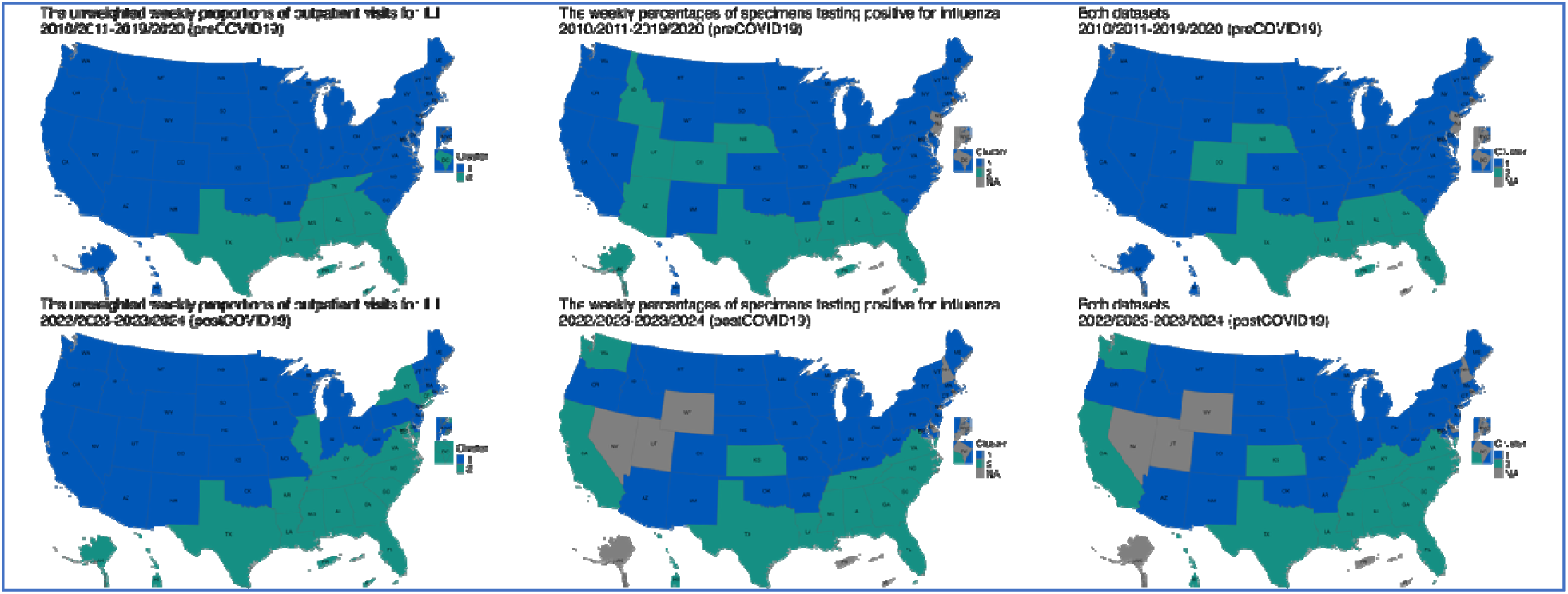
Spatial distribution of clustering patterns for the pre-COVID-19 seasons (2010/2011–2019/2020, upper panels) and the post-COVID-19 seasons (2022/2023–2023/2024, lower panels). (Left panels) Univariate analysis based on the unweighted weekly proportions of outpatient visits for ILI, (Middle panels) univariate analysis based on the weekly percentages of specimens testing positive for influenza, and (Right panels) multivariate analysis using both datasets. The blue color represents the largest cluster containing most jurisdictions, while the green color represents the smaller cluster with fewer jurisdictions. The gray jurisdictions were excluded from the analysis due to insufficient data across all seasons.

The two univariate analyses, based on different datasets (the unweighted weekly proportions of outpatient visits for ILI and the weekly percentages of specimens testing positive for influenza) presented slightly different patterns. The former included Tennessee (TN) in the Southeastern cluster, as well as the U.S. Virgin Islands (VI) and District of Columbia (DC), while the latter included some Central states such as Colorado (CO) and Nebraska (NE). In the multivariate analysis using both datasets, Colorado (CO) and Nebraska (NE) remained within the Southeastern cluster. We then verified cluster membership by identifying jurisdictions within one standard deviation from their respective centroids and found only a limited number of outliers (Figure S7). Notably, the clustering patterns of the 5 core Southeastern members remained robust during the post-COVID-19 seasons (2022/2023–2023/2024) (Figure 2).

Clustering patterns demonstrated inter-season variability throughout the study period (Figure 3, Figure S8 and Figure S9). While the core Southeastern members generally maintained consistent cluster associations, some other jurisdictions experienced significant season-to-season changes in cluster membership, highlighting the dynamic nature of influenza activity. The Western and Central states showed the greatest variability, with shifts between clusters from one season to the next.

**Figure 3.**
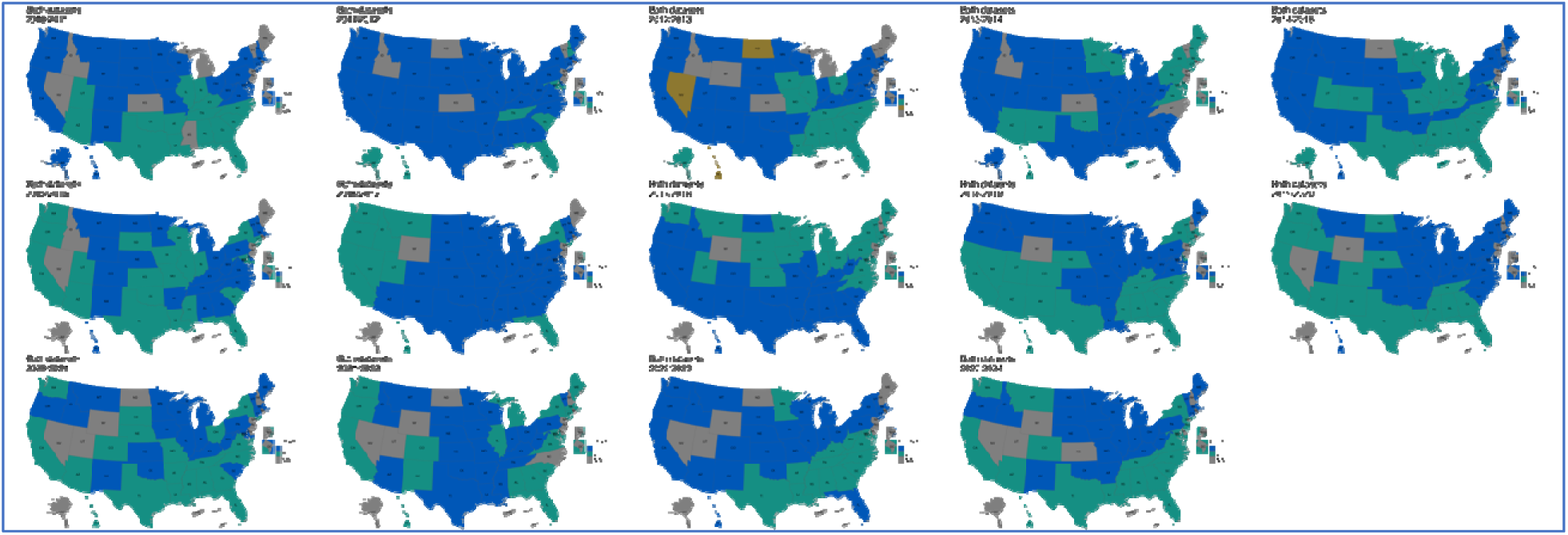
Spatial distribution of clustering patterns from the 2010/2011 to 2023/2024 seasons, based on multivariate analysis using both datasets (the unweighted weekly proportions of outpatient visits for ILI and the weekly percentages of specimens testing positive for influenza). The latter consist of both public health and clinical laboratory data reported before the 2015/2016 season, while only the clinical laboratory data were available from the 2015/2016 season onward. The blue color represents the largest cluster containing most jurisdictions, while the green color represents the smaller cluster with fewer jurisdictions. In the 2012/2013 season, three clusters were identified, represented by blue, green, and brown colors, corresponding to the largest, second smallest, and smallest clusters, respectively. The gray jurisdictions were excluded from the analysis due to insufficient data for the respective seasons.

### Significant differences across clusters in local Moran’s I and proportion of A/H1 viruses

During the pre-COVID-19 seasons (2010/2011–2019/2020), the ANOVA revealed significant differences across clusters in local Moran’s I (p < 0.001) and the proportion of all influenza A virus detections that were influenza A/H1 viruses (p < 0.001). The Tukey HSD test confirmed that the smaller cluster with fewer jurisdictions exhibited significantly higher spatial autocorrelation (mean difference in local Moran’s I = 0.16) compared to the larger cluster. This suggests that influenza activity within the smaller cluster (typically the Southeastern cluster) was more geographically synchronized. Similarly, the Tukey HSD test indicated that the smaller cluster had a significantly higher proportion of all influenza A virus detections that were influenza A/H1 viruses (mean difference = 2%) compared to the larger cluster.

Although the magnitude of the difference was relatively small, this indicates potential spatial heterogeneity in viral circulation across regions.

In contrast, we found no significant differences across clusters in peak timing (p = 0.825) or the proportion of all influenza A and B virus detections that were influenza A viruses (p = 0.0623). However, significant interactions between clusters and seasons indicated that temporal variability may also influence these relationships (Table S2). Notably, seasonal variation emerged as a dominant factor influencing all four dependent variables: peak timing, local Moran’s I, the proportion of all influenza A and B virus detections that were influenza A viruses, and the proportion of all influenza A virus detections that were influenza A/H1 viruses (all p < 0.001). This suggests that seasonal variability in influenza activity played a key role in shaping spatial patterns and aligns with the inter-season variability identified in earlier sections. Further details are provided in the supplementary materials (Figure S10, Figure S11, Figure S12, Figure S13, and Table S2).

## Discussion

### Consistent spatial clustering patterns of seasonal influenza

Our study reveals distinct spatial clustering patterns of seasonal influenza in the United States. We found a consistent grouping of Southeastern states, particularly Georgia, Alabama, Mississippi, Louisiana, and Florida, suggesting a specific spatiotemporal pattern of seasonal influenza for this region that was also reflected in earlier seasonal peaks and high local spatial autocorrelation.

Our findings highlight regional variations in influenza dynamics that could have implications for public health strategies. The consistent clustering observed in Southeastern states suggests that these areas tend to experience earlier influenza activity on average. Recognizing these patterns can help inform influenza prevention and control efforts, including timing of vaccination campaigns and public communication. Furthermore, the identification of outliers within clusters, and the occasional inclusion of Western or Central states with the Southeastern cluster, emphasizes the importance of considering local factors that influence influenza activity. Maintaining robust surveillance and long-term monitoring across regions should provide valuable insights for refining broader public health responses to seasonal influenza.

### Seasonal variations

The variation in clustering patterns observed across individual seasons underscores the dynamic nature of influenza activity. These season-to-season variations, as seen in the significant interactions between clusters and seasons, highlight the inherent challenges in forecasting influenza trends. Our findings closely align with the earlier work of Rosensteel et al. (Rosensteel, et al. 2021), who characterized significant heterogeneity in influenza activity across seasons during the 2002–2009 period, and Dahlgren et al. (Dahlgren, et al. 2019), who found substantial variation during the 2010–2016 seasons. Our results demonstrate that this spatiotemporal heterogeneity continued in the post-2009 pandemic era and following the COVID-19 pandemic, e.g., in the 2022/2023 and 2023/2024 seasons.

Longer-term analyses are needed to fully understand the impact of COVID-19 pandemic on influenza activity patterns. These disruptions are likely due to changes in population behavior, non-pharmaceutical interventions, and reduced exposure to influenza viruses during the period. For instance, the impact of school openings on influenza spread has been well-documented, particularly during the 2009 pandemic (Gog, et al. 2014). Additionally, the emergence of antigenically drifted influenza viruses and changes in genetic clades can further alter epidemic patterns and severity.

Integrating such insights with longer time series data could provide a more comprehensive understanding of the shifts observed during and after the COVID-19 pandemic.

### Data variability

Our analysis revealed some variations in cluster membership by data source, particularly for the Western and Central states, which occasionally showed similarities to the Southeastern cluster when using the weekly percentages of specimens testing positive for influenza. The data on specimens testing positive for influenza virus were limited by missing values, particularly during off-seasons, which may affect the robustness of our analysis for some jurisdictions. Additionally, outpatient visits for ILI lack laboratory confirmation, and may therefore capture activity of other respiratory viruses in addition to influenza-specific activity.

Despite these challenges, our univariate analyses provide unique insights from each data source. For example, using the unweighted weekly proportions of outpatient visits for ILI allowed us to explore patterns for the U.S. Virgin Islands and the District of Columbia, even though these were not included in the weekly percentages of specimens testing positive for influenza virus. We also performed a multivariate approach to combine information from both datasets and confirm the identification of the Southeastern cluster. However, as these data are based on healthcare visits and testing, there may still be potential biases related to healthcare access and care-seeking behaviors.

Our findings highlight the potential influence of factors beyond geographic proximity on spatial patterns of influenza activity. Additional data sources including climate variables, environmental factors, population density, and socioeconomic indicators may help explain observed deviations from spatial clustering. For example, earlier studies (Shaman and Kohn 2009) (Shaman, Pitzer, et al. 2010) demonstrated that humidity plays a key role in seasonal influenza transmission and Dalziel et al. (Dalziel, et al. 2018) further highlighted that humidity is a significant driver particularly in urban areas. Extending these works to incorporate temperature, air quality, and other environmental variables could offer a broader understanding of their influence on influenza transmission. Recent work by Chen et al. (Chen, et al. 2024) found positive correlations between influenza case counts and several air pollutants in China, suggesting a need to explore similar environmental interactions in the United States.

### State and territory level heterogeneity

Our analysis was limited to state and territory levels, which may mask finer-scale heterogeneity in influenza activity patterns. County-level analysis, such as in Rosensteel et al. (Rosensteel, et al. 2021), and studies of core-based statistical areas, such as Dahlgren et al. (Dahlgren, et al. 2019), could reveal additional important patterns, especially in densely populated urban areas. One key factor contributing to such finer-scale heterogeneity is human mobility. While state and territory level analyses offer broad regional insights, influenza transmission is heavily influenced by movement patterns within and between cities, commuting zones, and rural areas (Viboud, et al. 2006) (Charu, et al. 2017). Future studies integrating mobility data, such as commuting flows and other travel behaviors, and examining their effects in both urban and rural settings, could enhance our understanding of spatial transmission patterns.

## Conclusions

This study quantified distinct spatial differences in seasonal influenza activity across the United States. Our results suggest that interpreting national influenza trends may overlook important regional variations. For example, the Southeastern United States, including Georgia, Alabama, Mississippi, Louisiana, and Florida, often experienced earlier influenza activity compared to other regions, although these spatiotemporal patterns were not consistent across seasons, reflecting the dynamic nature of influenza transmission. More generally, our findings emphasize the importance of robust state-based influenza surveillance systems, like those supported by CDC, to provide actionable information for understanding regional influenza dynamics. Adaptive approaches and stable long-term surveillance data are critical for effectively addressing regional differences in influenza activity. The potential impacts of variation in seasonal timing should be considered when planning influenza prevention and control efforts, including timing of vaccination campaigns and public communication, to enhance preparedness and effectiveness nationwide.

## Data Availability

The surveillance data utilized in this study are publicly available through the CDC's FluView Interactive platform (https://gis.cdc.gov/grasp/fluview/fluportaldashboard.html).

https://github.com/CDCgov/influenza-cluster-us

https://gis.cdc.gov/grasp/fluview/fluportaldashboard.html

## Declarations

### Ethics approval and consent to participate

Not applicable.

### Consent for publication

Not applicable.

### Availability of data and materials

The surveillance data utilized in this study are publicly available through the CDC’s FluView Interactive platform (https://gis.cdc.gov/grasp/fluview/fluportaldashboard.html). All analyses were performed using R version 4.4.0. The R scripts and code supporting the findings of this study are accessible on GitHub at https://github.com/CDCgov/influenza-cluster-us.

### Authors’ contributions

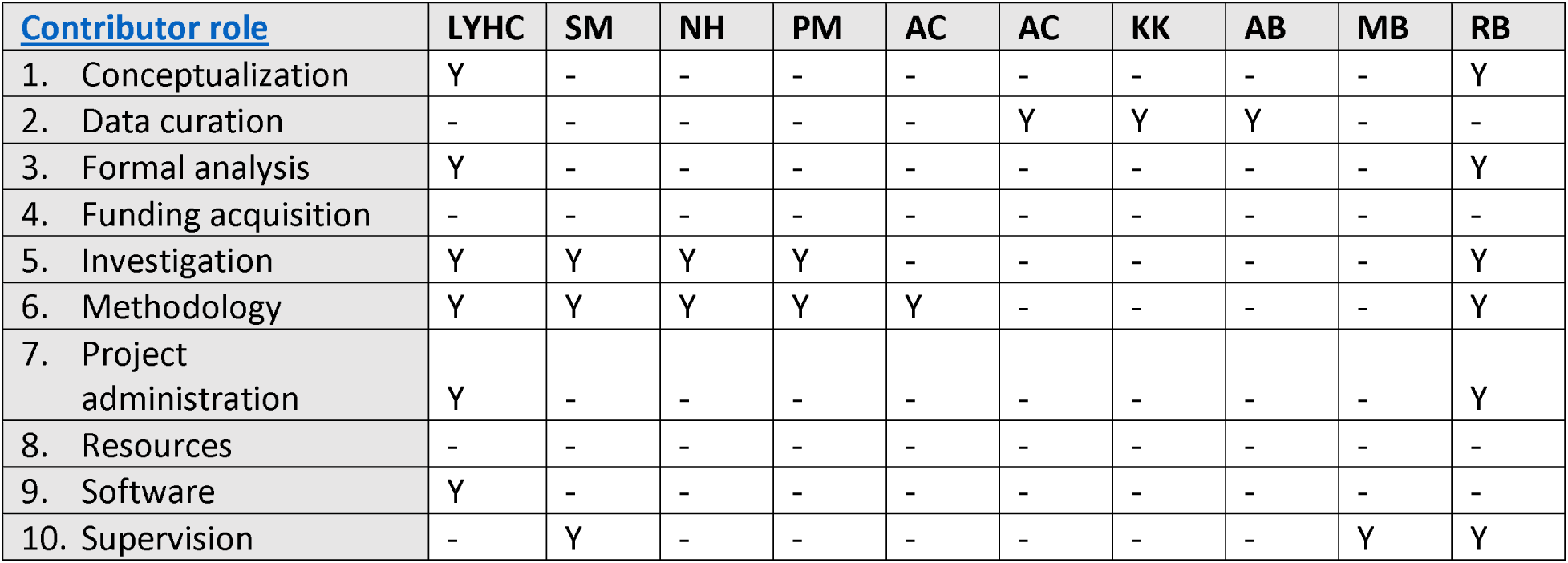
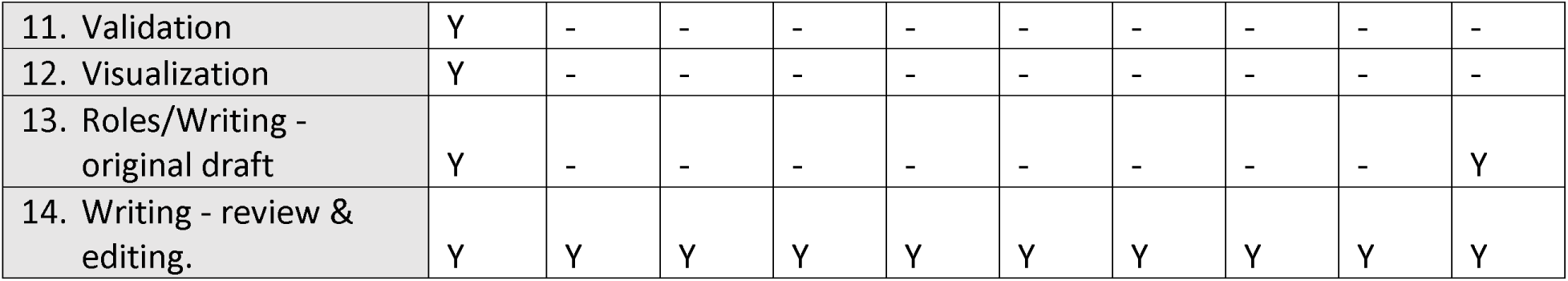

### Statements of contributions

- L.Y.H. Chan contributed to conceptualization, formal analysis, investigation, methodology, project administration, software, validation, visualization, drafting the original manuscript, and reviewing and editing the manuscript.
- S. Morris contributed to investigation, methodology, supervision, and reviewing and editing the manuscript.
- N. Hassell contributed to investigation, methodology, and reviewing and editing the manuscript.
- P. Marcenac contributed to investigation, methodology, and reviewing and editing the manuscript.
- A. Couture contributed to methodology and reviewing and editing the manuscript.
- A. Colon contributed to data curation and reviewing and editing the manuscript.
- K. Kniss contributed to data curation and reviewing and editing the manuscript.
- A. Budd contributed to data curation and reviewing and editing the manuscript.
- M. Biggerstaff contributed to supervision and reviewing and editing the manuscript.
- R. Borchering contributed to conceptualization, formal analysis, investigation, methodology, project administration, supervision, drafting the original manuscript, and reviewing and editing the manuscript.
- All authors reviewed and approved the final manuscript.

## Acknowledgements

The authors thank the state, county, city, and territorial health departments and public health laboratories for collecting and reporting the surveillance data.

• We also thank the CDC surveillance teams for processing and maintaining the data on FluView Interactive.

• L.Y.H. Chan thanks the CDC Steven M. Teutsch Prevention Effectiveness (PE) Fellowship.

## Authors’ information

L.Y.H. Chan, N. Hassell, P. Marcenac, A. Couture, A. Colon, K. Kniss, A. Budd, M. Biggerstaff, and R. Borchering are employees of the CDC.

## Disclaimer

The conclusions, findings, and opinions expressed by authors contributing to this article do not necessarily reflect the official position of the U.S. Department of Health and Human Services, the Public Health Service, the Centers for Disease Control and Prevention, or the authors’ affiliated institutions.

## Declaration of using large language models in the writing process

During the preparation of this work the authors used *CDC AI Chatbot (GPT-4o)* in order to enhance the clarity, coherence, and correctness of the writing, and to check for grammatical errors. After using this tool, the authors reviewed and edited the content as needed and take full responsibility for the content of the publication.

## Supplementary material

### Surveillance data

For the 2020/2021–2023/2024 seasons, data availability differed from the pre-COVID-19 period (Figure S1). The unweighted weekly proportions of outpatient visits for ILI were available for all jurisdictions. However, the weekly percentages of specimens testing positive for influenza included fewer jurisdictions compared to the pre-COVID-19 seasons, though the jurisdictions included in the analysis exhibited fewer missing data (Table S1). Importantly, influenza activity levels during the 2020/2021 season were notably lower, reflecting the widespread impact of the COVID-19 pandemic.

**Table S1.** Number of jurisdictions included in the analysis after data processing, along with the proportion of available data for the jurisdictions included.

| Study period | The unweighted weekly proportions of outpatient visits for ILI |  | The weekly percentages of specimens testing positive for influenza |  |
| --- | --- | --- | --- | --- |
|  | Number of jurisdictions | Proportion of available data | Number of jurisdictions | Proportion of available data |
| **pre-COVID-19 season** |  |  |  |  |
| 2010/2011 | 52 | 1.00 | 41 | 0.87 |
| 2011/2012 | 53 | 1.00 | 44 | 0.87 |
| 2012/2013 | 53 | 1.00 | 42 | 0.89 |
| 2013/2014 | 54 | 1.00 | 45 | 0.90 |
| 2014/2015 | 54 | 1.00 | 47 | 0.94 |
| 2015/2016 | 54 | 1.00 | 44 | 0.96 |
| 2016/2017 | 54 | 1.00 | 45 | 0.95 |
| 2017/2018 | 54 | 1.00 | 45 | 0.96 |
| 2018/2019 | 54 | 1.00 | 45 | 0.97 |
| 2019/2020 | 54 | 1.00 | 44 | 0.96 |
| **COVID-19-era season** |  |  |  |  |
| 2020/2021 | 54 | 1.00 | 41 | 0.98 |
| 2021/2022 | 54 | 1.00 | 40 | 0.98 |
| **post-COVID-19 season** |  |  |  |  |
| 2022/2023 | 54 | 1.00 | 40 | 0.99 |
| 2023/2024 | 54 | 1.00 | 42 | 0.99 |

**Figure S1.**
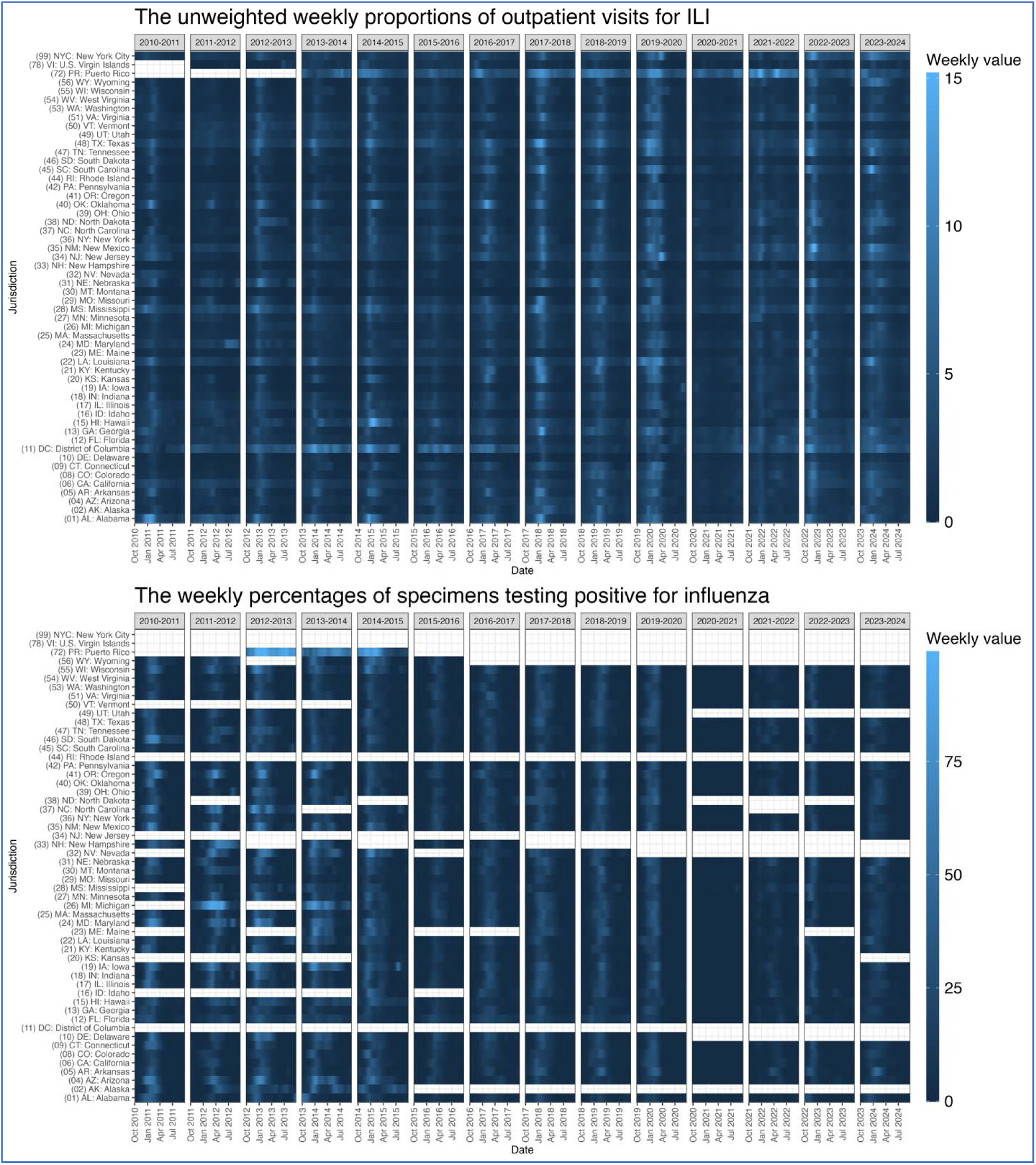
Weekly values of the unweighted weekly proportions of outpatient visits for ILI and the weekly percentages of specimens testing positive for influenza. The latter consist of both public health and clinical laboratory data reported before the 2015/2016 season, while only the clinical laboratory data were available from the 2015/2016 season onward. All weekly values were processed by imputation and smoothing. The color scale ranges from dark to light blue, indicating values from low to high. White space represents jurisdictions with more than 50% missing data, which were excluded from the analysis due to insufficient data for the respective seasons.

### Peak timing and Moran’s I

**Figure S2.**
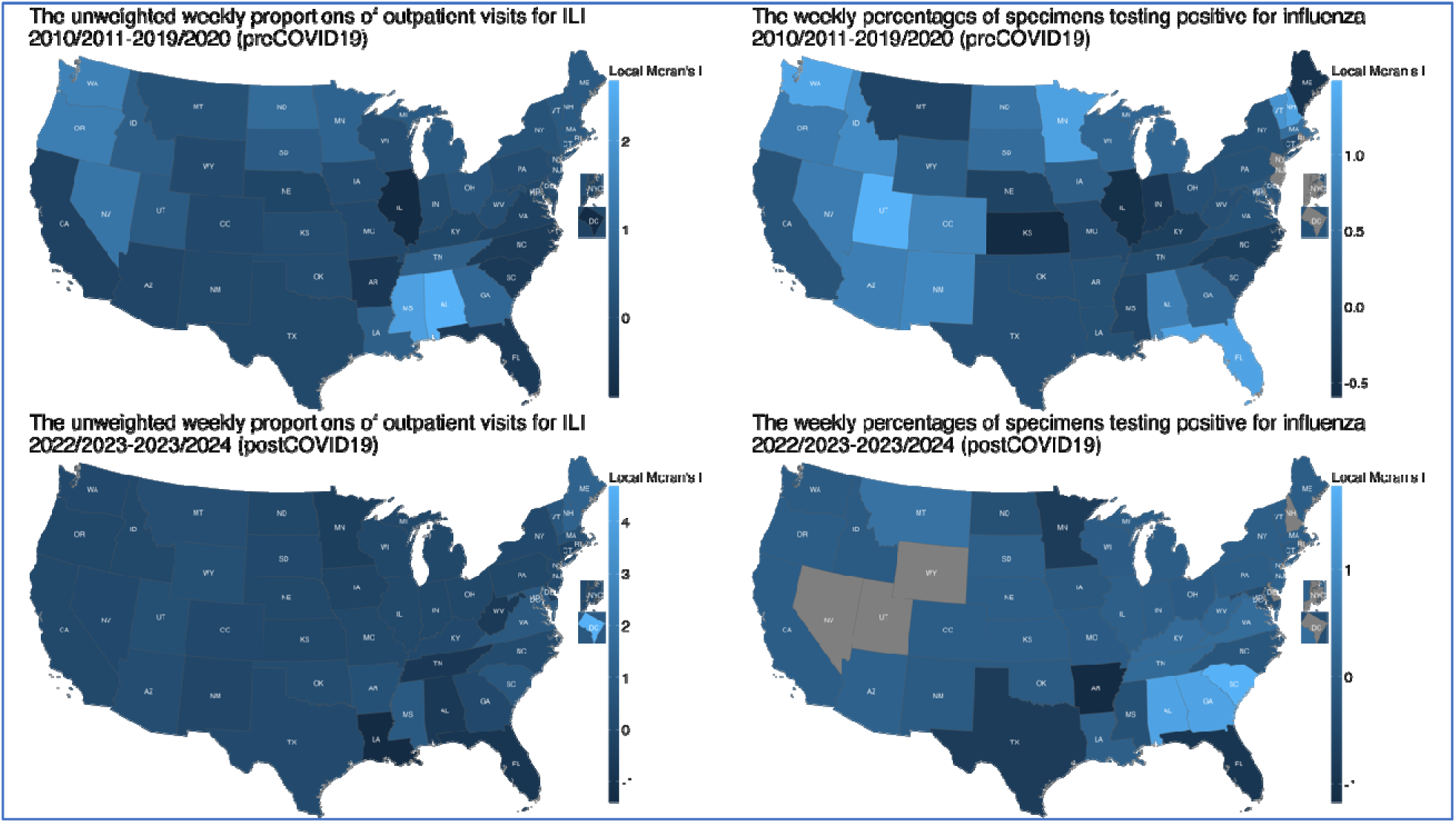
Spatial distribution of local Moran’s I values for the unweighted weekly proportions of outpatient visits for ILI (left panels) and the weekly percentages of specimens testing positive for influenza (right panels), averaged across the pre-COVID-19 seasons (2010/2011–2019/2020, upper panels) and the post-COVID-19 seasons (2022/2023–2023/2024, lower panels). The latter consist of both public health and clinical laboratory data reported before the 2015/2016 season, while only the clinical laboratory data were available from the 2015/2016 season onward. Lighter blue indicates higher local Moran’s I values (stronger clustering), while darker blue represents lower values. The gray color (e.g., in DC, NJ, and NYC) indicates that all seasons were excluded from the analysis due to missing data.

**Figure S3.**
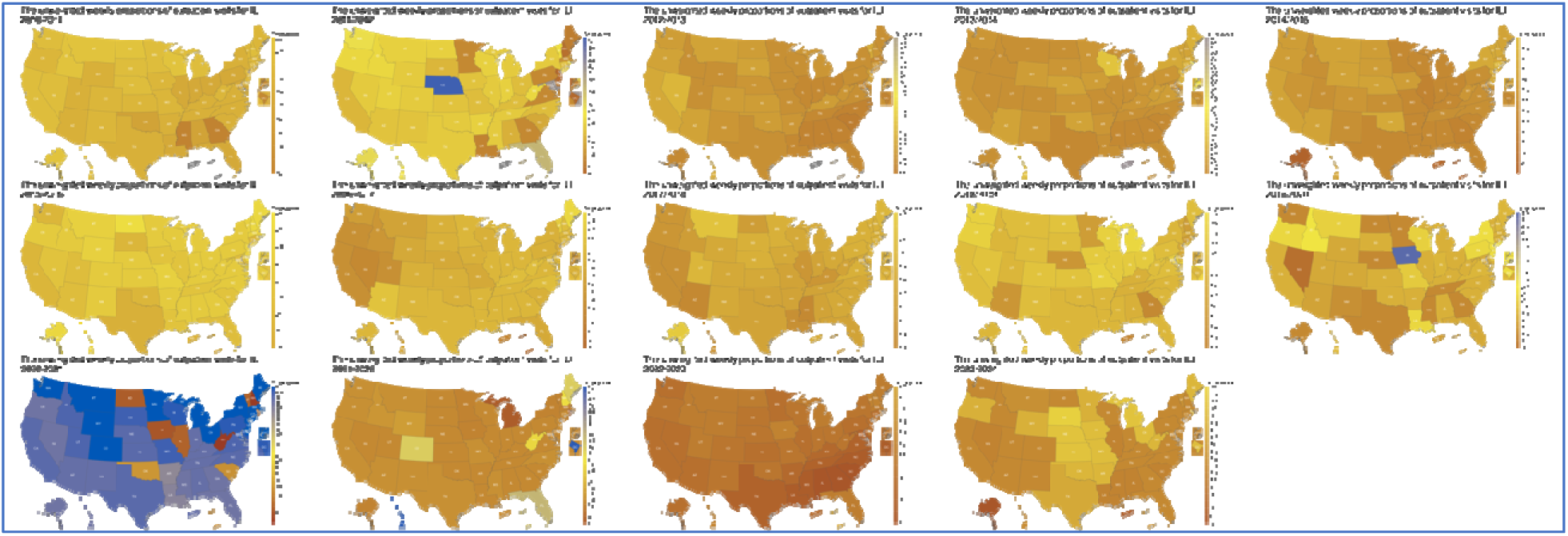
Spatial distribution of peak timing for the unweighted weekly proportions of outpatient visits for ILI, in each season (2010/2011–2023/2024). Darker yellow indicates earlier peak timing, while lighter yellow represents later peak timing. All panels share the same color scale, which corresponds to the week from the beginning of influenza season (MMWR Week 40). The gray color indicates that jurisdictions were excluded from the analysis due to missing data for the respective seasons.

**Figure S4.**
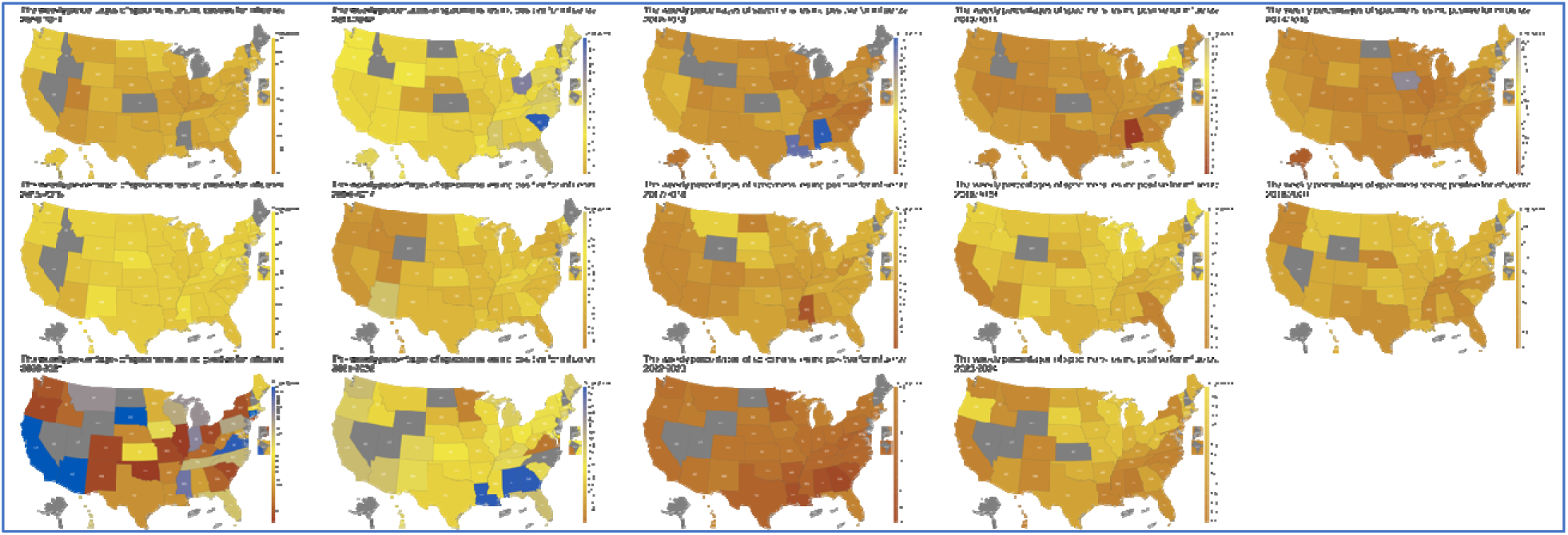
Spatial distribution of peak timing for the weekly percentages of specimens testing positive for influenza, in each season (2010/2011–2023/2024). The data consist of both public health and clinical laboratory data reported before the 2015/2016 season, while only the clinical laboratory data were available from the 2015/2016 season onward. Darker yellow indicates earlier peak timing, while lighter yellow represents later peak timing. All panels share the same color scale, which corresponds to the week from the beginning of influenza season (MMWR Week 40). The gray color indicates that jurisdictions were excluded from the analysis due to missing data for the respective seasons.

**Figure S5.**
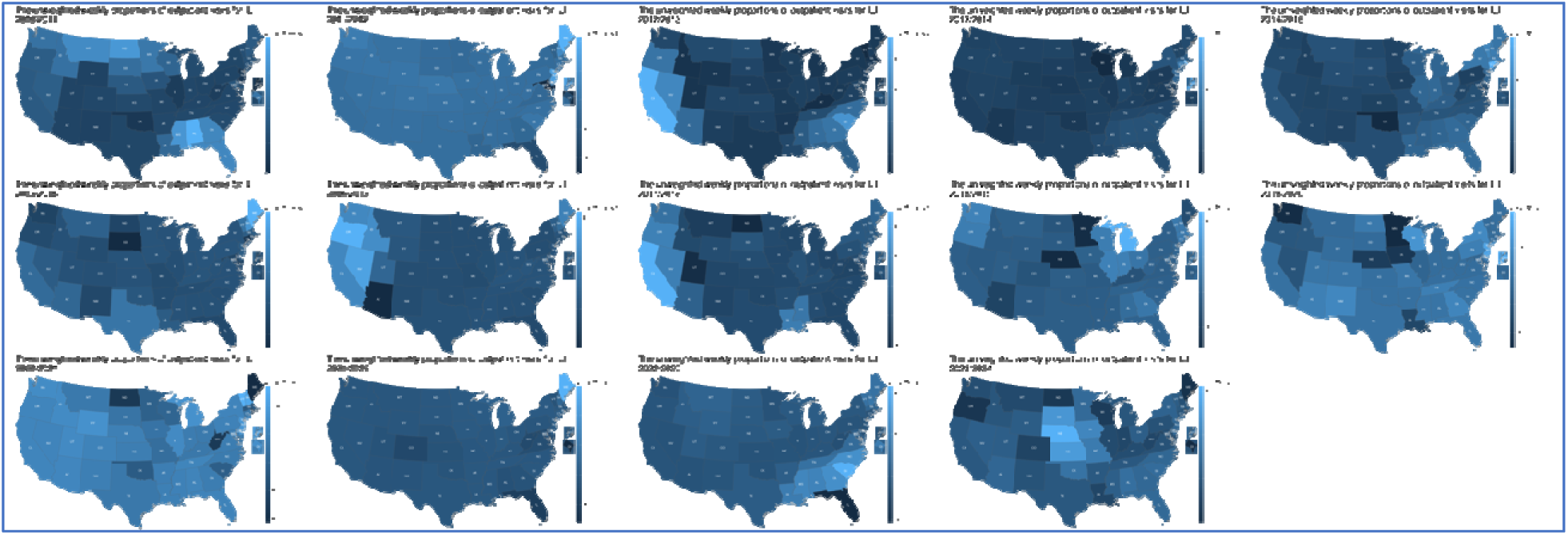
Spatial distribution of local Moran’s I values for the unweighted weekly proportions of outpatient visits for ILI, in each season (2010/2011–2023/2024). Lighter blue indicates higher local Moran’s I values (stronger clustering), while darker blue represents lower values. The gray color indicates that jurisdictions were excluded from the analysis due to missing data for the respective seasons.

**Figure S6.**
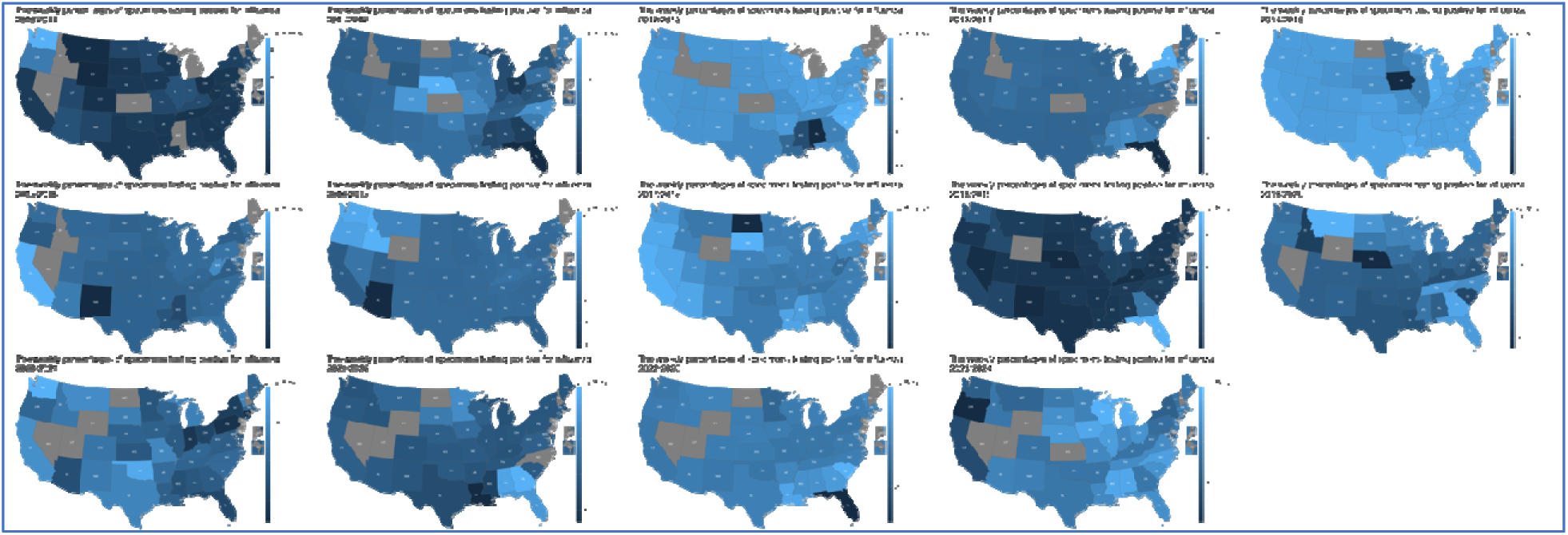
Spatial distribution of local Moran’s I values for the weekly percentages of specimens testing positive for influenza, in each season (2010/2011–2023/2024). The data consist of both public health and clinical laboratory data reported before the 2015/2016 season, while only the clinical laboratory data were available from the 2015/2016 season onward. Lighter blue indicates higher local Moran’s I values (stronger clustering), while darker blue represents lower values. The gray color indicates that jurisdictions were excluded from the analysis due to missing data for the respective seasons.

### Spatial clustering patterns

**Figure S7.**
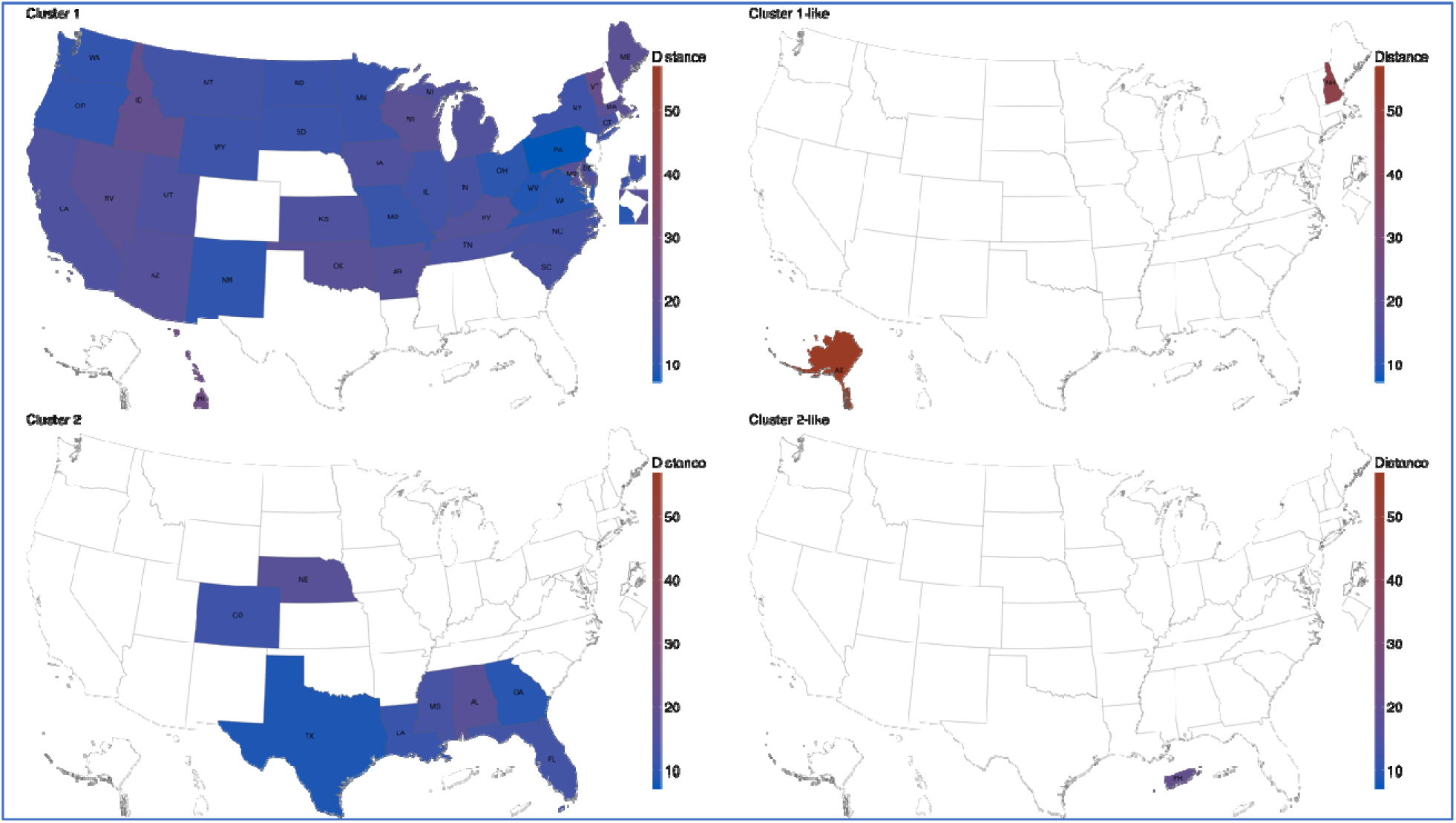
Spatial distribution of cluster distances from their respective centroids, based on the multivariate analysis using both datasets (the unweighted weekly proportions of outpatient visits for ILI and the weekly percentages of specimens testing positive for influenza) for the pre-COVID-19 seasons (2010/2011–2019/2020, upper right panel of Figure 2). The latter consist of both public health and clinical laboratory data reported before the 2015/2016 season, while only the clinical laboratory data were available from the 2015/2016 season onward. The blue color represents shorter distances, while red represents longer distances. The right panels of this figure show outliers, defined as those with a distance greater than one standard deviation from the mean. New Hampshire (NH) and Alaska (AK) are outliers in Cluster 1, while Puerto Rico (PR) is the outlier in Cluster 2.

**Figure S8.**
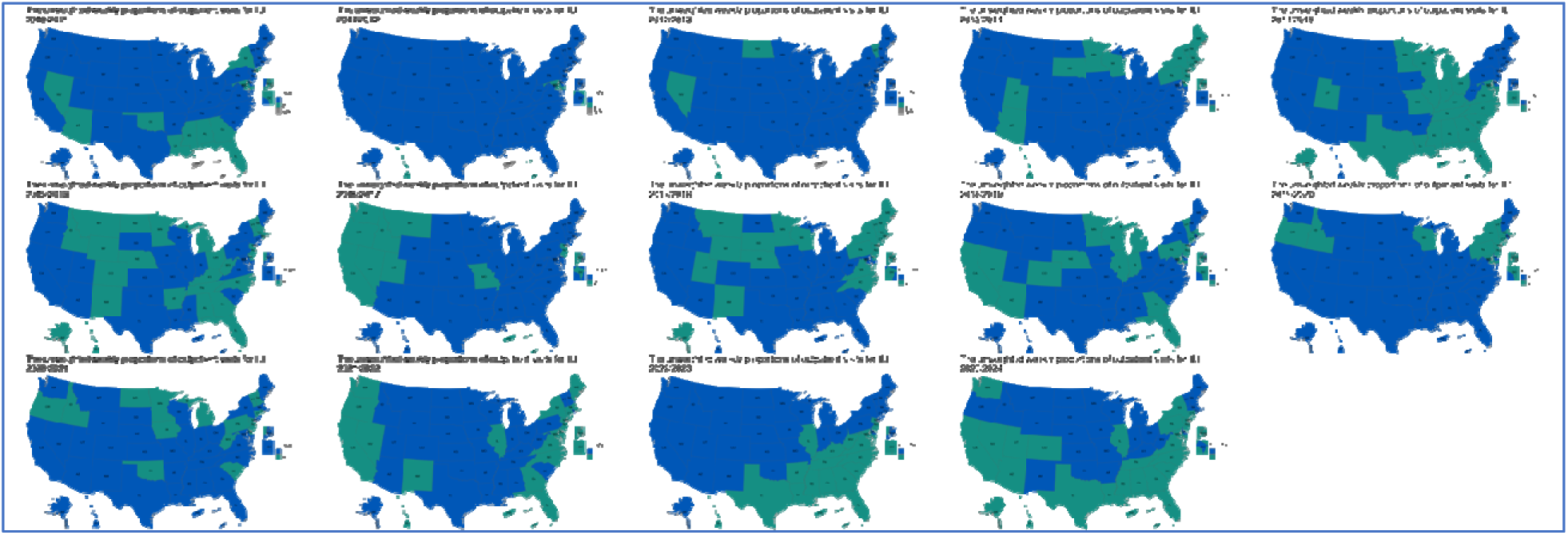
Spatial distribution of clustering patterns from the 2010/2011 to 2023/2024 seasons, based on univariate analysis using the unweighted weekly proportions of outpatient visits for ILI. The blue color represents the largest cluster containing most jurisdictions, while the green color represents the smaller cluster with fewer jurisdictions. The gray jurisdictions were excluded from the analysis due to insufficient data for the respective seasons.

**Figure S9.**
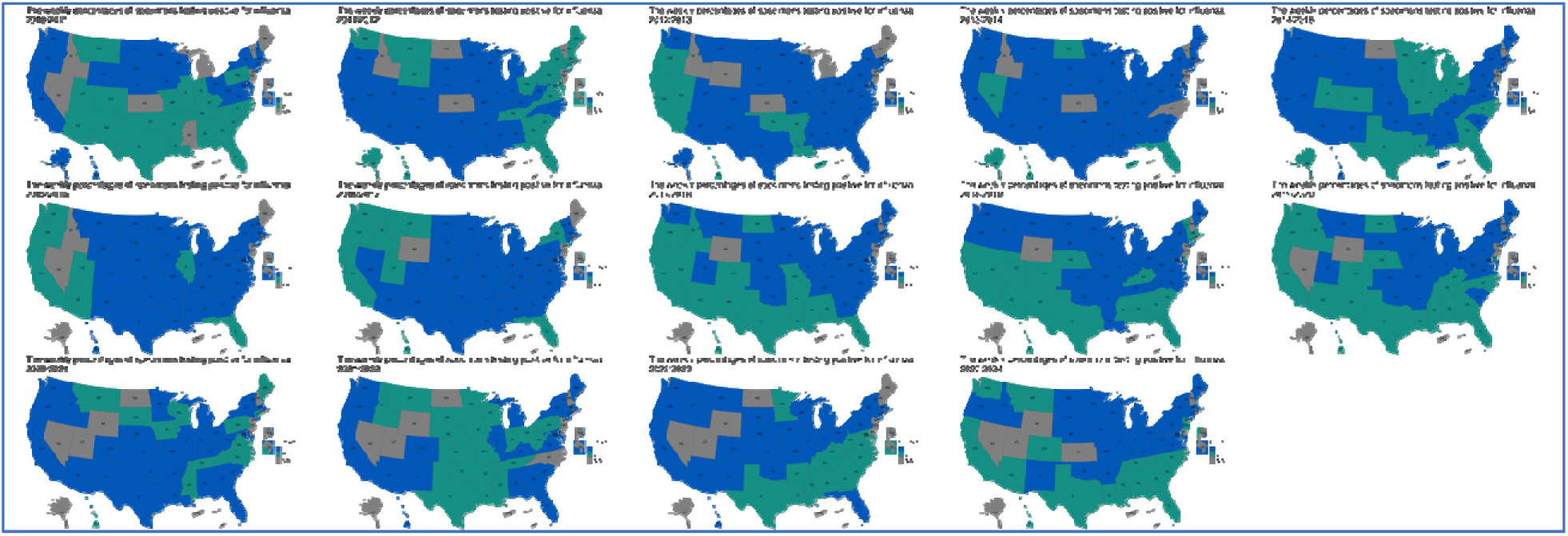
Spatial distribution of clustering patterns from the 2010/2011 to 2023/2024 seasons, based on univariate analysis using the weekly percentages of specimens testing positive for influenza. The data consist of both public health and clinical laboratory data reported before the 2015/2016 season, while only the clinical laboratory data were available from the 2015/2016 season onward. The blue color represents the largest cluster containing most jurisdictions, while the green color represents the smaller cluster with fewer jurisdictions. The gray jurisdictions were excluded from the analysis due to insufficient data for the respective seasons.

### Evaluating cluster differences using ANOVA

We conducted a series of three-way ANOVAs to examine the effects of CLUSTER (with values 1, 2, or 3), SEASON (10 seasons: 2010/2011–2019/2020), and DATASET (the unweighted weekly proportions of outpatient visits for ILI, the weekly percentages of specimens testing positive for influenza, or both) on four key variables: PEAK_WEEK (peak timing), MORAN_I_LOCAL (local Moran’s I), PERCENT_A (proportion of all influenza A and B virus detections that were influenza A viruses), and PERCENT_H1 (proportion of all influenza A virus detections that were influenza A/H1 viruses). The distributions of each of these variables are shown in Figure S10, Figure S11, Figure S12 and Figure S13. We note that only univariate analyses were performed for PEAK_WEEK and MORAN_I_LOCAL, and the optimal number of clusters was .

The ANOVA summary is provided in Table S2.

- PEAK_WEEK (peak timing): Significant differences were found for SEASON, CLUSTER:SEASON, CLUSTER:DATASET, and CLUSTER:SEASON:DATASET interactions. This indicates that peak timing varied across seasons, between clusters, and according to the data type, suggesting complex seasonal patterns for influenza.
- MORAN_I_LOCAL (local Moran’s I): Significant effects were observed for CLUSTER, SEASON, DATASET, and their interactions (except CLUSTER:DATASET). This suggests that local spatial autocorrelation patterns were influenced by clustering, seasonal variations, and data types, with distinct spatial patterns over time.
- PERCENT_A (proportion of Influenza A viruses): Significant variation was observed for SEASON and the CLUSTER:SEASON interaction. This indicates that the distribution of Influenza A varied across seasons and clusters.
- PERCENT_H1 (proportion of Influenza A/H1 viruses): Significant effects were found for CLUSTER, SEASON, and the CLUSTER:SEASON:DATASET interaction. This shows that the proportion of A/H1 viruses differed between clusters, varied over seasons, and was influenced by the complex interaction of all three factors.

**Figure S10.**
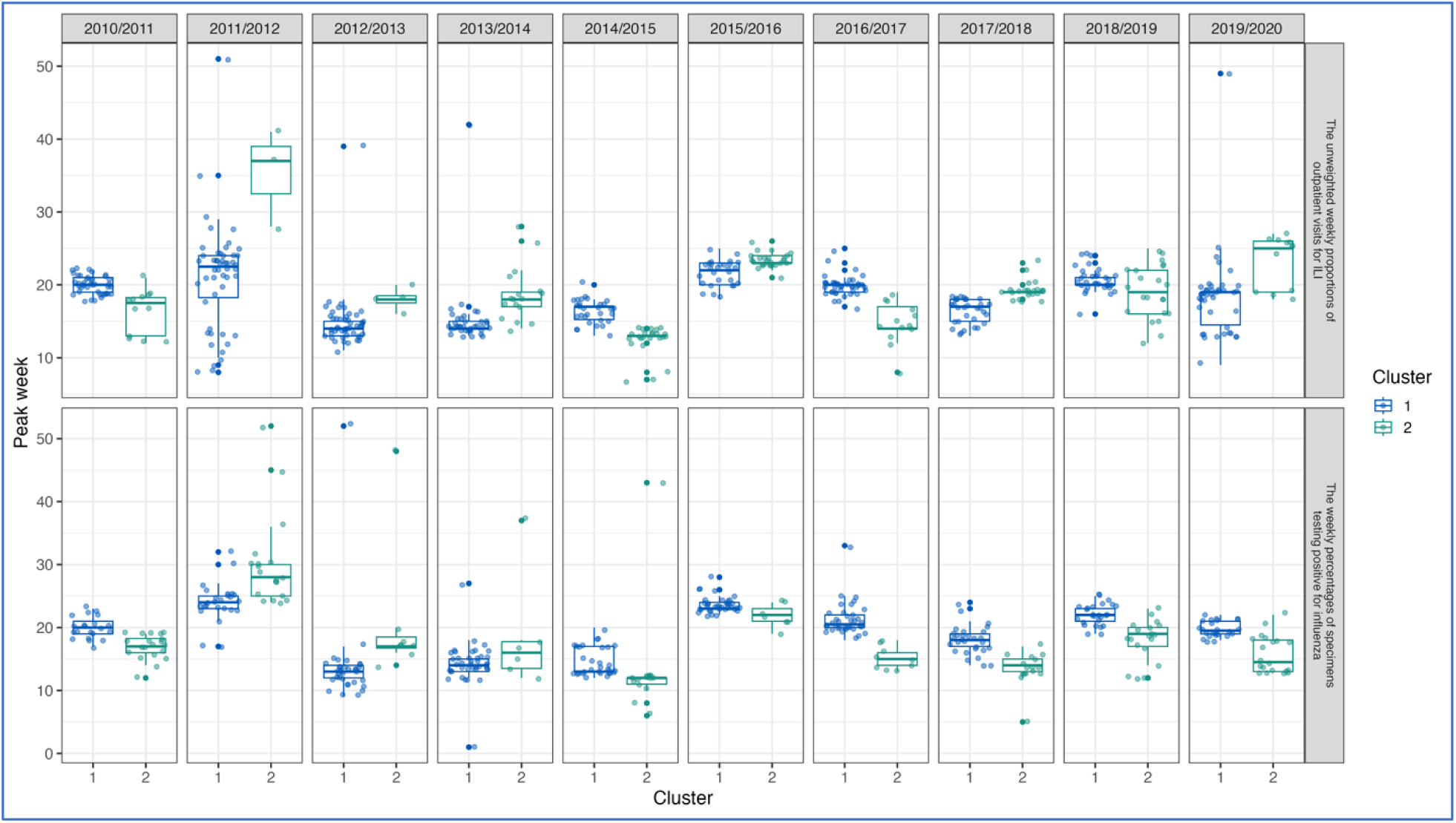
Distributions of peak timing across clusters, seasons and datasets. Only univariate analyses were performed.

**Figure S11.**
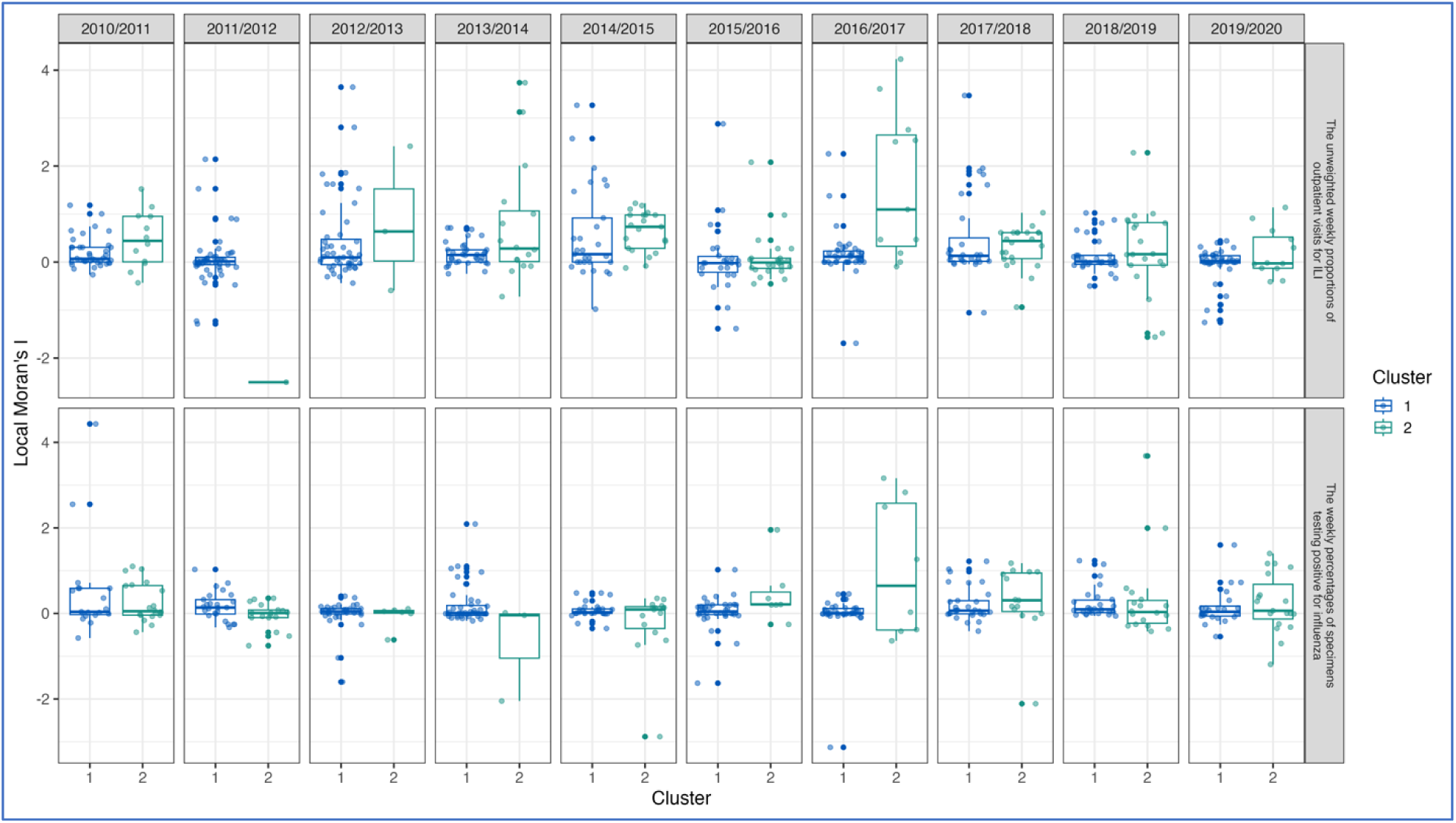
Distributions of local Moran’s I across clusters, seasons and datasets. Only univariate analyses were performed.

**Figure S12.**
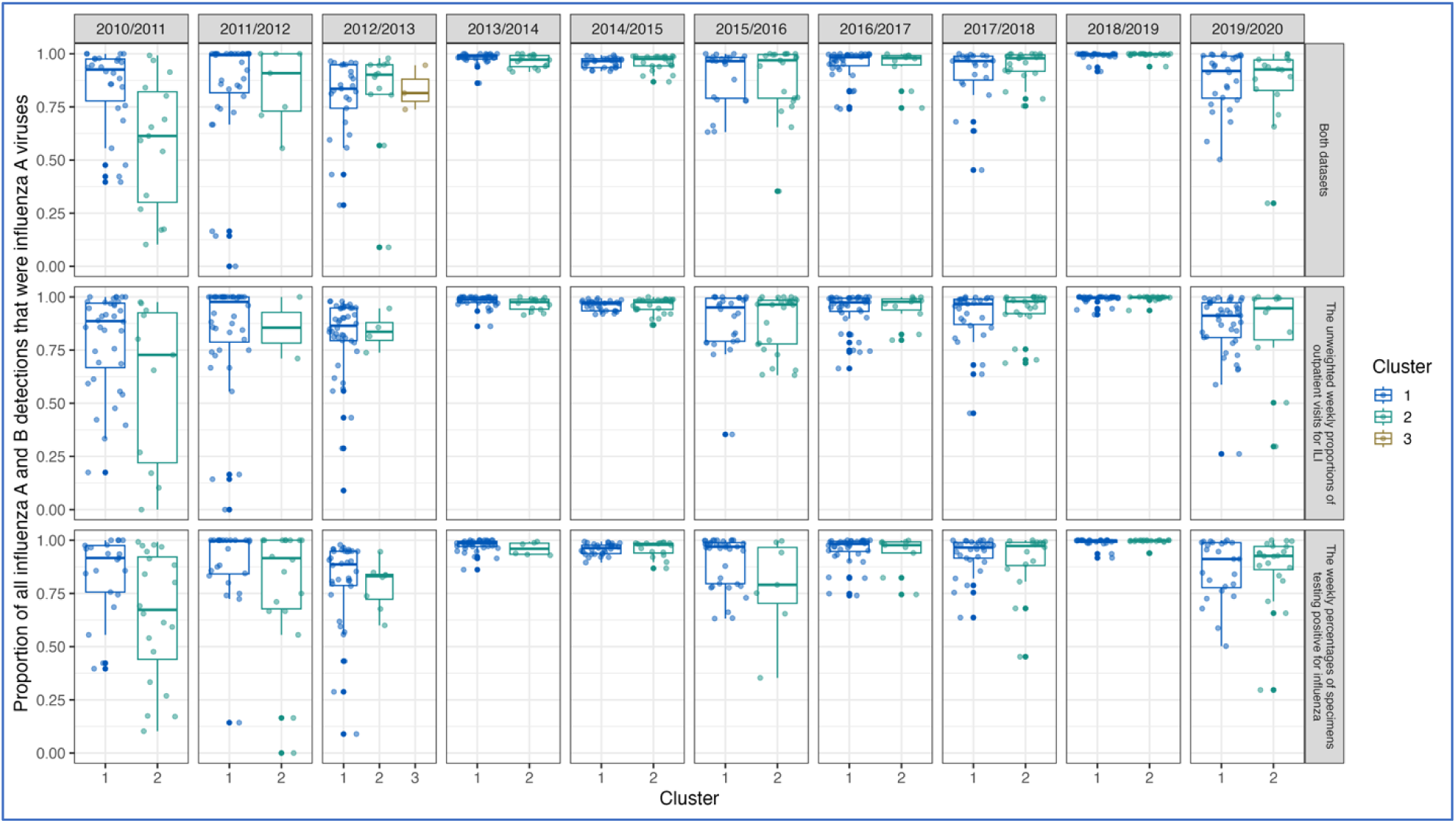
Distributions of the proportion of all influenza A and B virus detections that were influenza A viruses across clusters, seasons and datasets.

**Figure S13.**
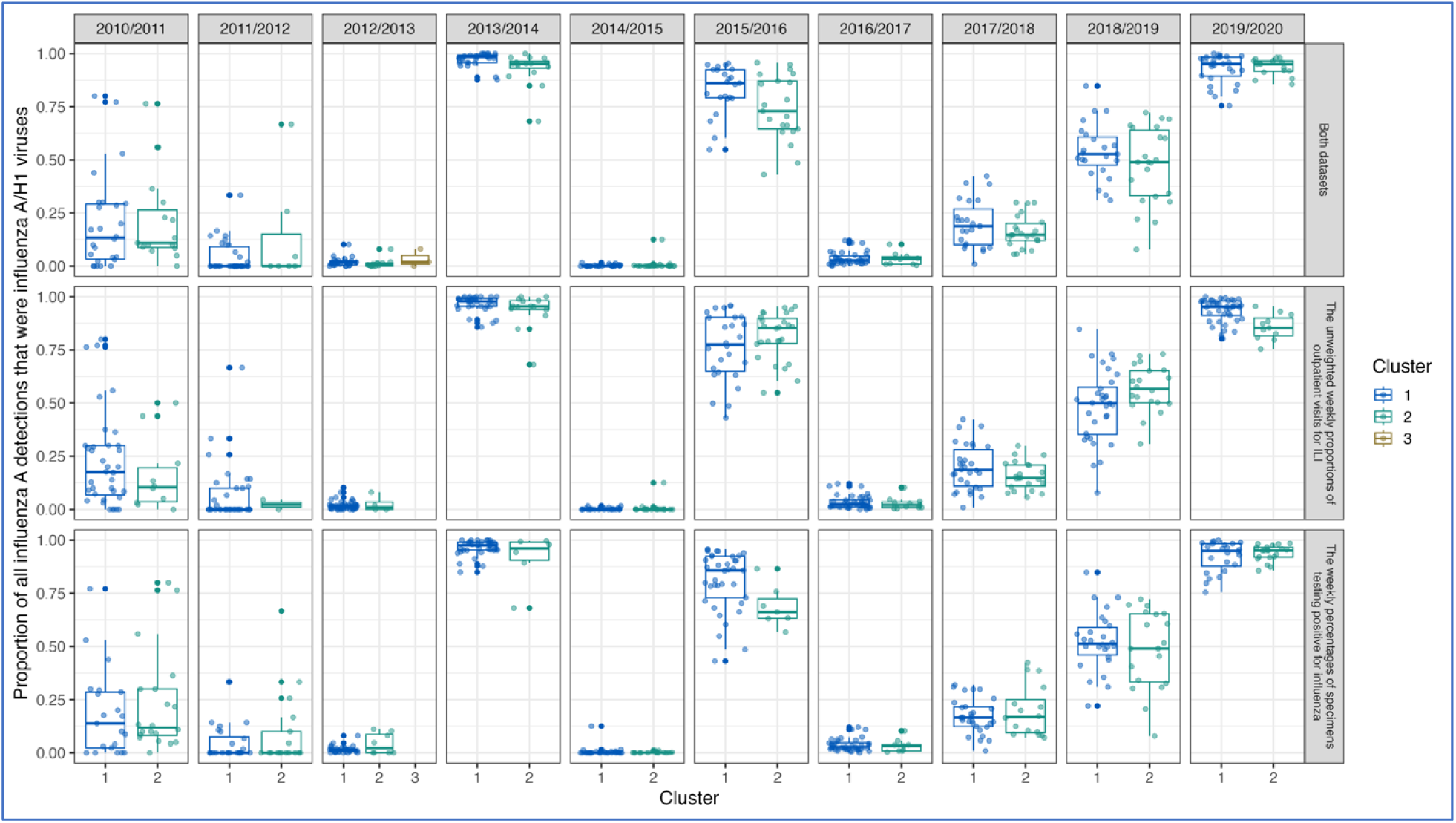
Distributions of the proportion of all influenza A virus detections that were influenza A/H1 viruses across clusters, seasons and datasets.

**Table S2.** Results of three-way ANOVAs examining the effects of CLUSTER (with values 1, 2, or 3), SEASON (10 seasons: 2010/2011–2019/2020), and DATASET (the unweighted weekly proportions of outpatient visits for ILI, the weekly percentages of specimens testing positive for influenza, or both) on four key variables: PEAK_WEEK (peak timing), MORAN_I_LOCAL (local Moran’s I), PERCENT_A (proportion of all influenza A and B virus detections that were influenza A viruses), and PERCENT_H1 (proportion of all influenza A virus detections that were influenza A/H1 viruses). Each column presents degrees of freedom (Df), sum of squares (Sum Sq), mean squares (Mean Sq), F-values, and p-values (Pr(>F)) for main effects and interactions. Significance levels are indicated as: *** p < 0.001, ** p < 0.01, * p < 0.05. Observations deleted due to missingness were 642 (PEAK_WEEK), 696 (MORAN_I_LOCAL), 245 (PERCENT_A), and 249 (PERCENT_H1).

| Source of Variation | Df | Sum Sq | Mean Sq | F value | Pr(>F) |
| --- | --- | --- | --- | --- | --- |
| **PEAK_WEEK** (peak timing) |  |  |  |  |  |
| CLUSTER | 1 | 1 | 0.7 | 0.049 | 0.825 |
| SEASON | 9 | 8997 | 999.6 | 67.175 | < 2e-16 *** |
| DATASET | 1 | 10 | 10.5 | 0.703 | 0.402 |
| CLUSTER:SEASON | 9 | 3007 | 334.1 | 22.453 | < 2e-16 *** |
| CLUSTER:DATASET | 1 | 306 | 305.6 | 20.534 | 6.61e-06 *** |
| SEASON:DATASET | 9 | 211 | 23.5 | 1.578 | 0.117 |
| CLUSTER:SEASON:DATASET | 9 | 758 | 84.2 | 5.661 | 1.18e-07 *** |
| Residuals | 938 | 13958 | 14.9 |  |  |
| **MORAN_I_LOCAL** (local Moran's I) |  |  |  |  |  |
| CLUSTER | 1 | 4.8 | 4.841 | 11.865 | 0.000599 *** |
| SEASON | 9 | 10.3 | 1.142 | 2.8 | 0.003036 ** |
| DATASET | 1 | 4.1 | 4.06 | 9.951 | 0.001662 ** |
| CLUSTER:SEASON | 9 | 24 | 2.669 | 6.542 | 4.58e-09 *** |
| CLUSTER:DATASET | 1 | 0.7 | 0.711 | 1.742 | 0.187263 |
| SEASON:DATASET | 9 | 17 | 1.886 | 4.622 | 5.41e-06 *** |
| CLUSTER:SEASON:DATASET | 9 | 13 | 1.441 | 3.531 | 0.000258 *** |
| Residuals | 884 | 360.7 | 0.408 |  |  |
| **PERCENT_A** (proportion of all influenza A and B virus detections that were influenza A viruses) |  |  |  |  |  |
| CLUSTER | 2 | 0.12 | 0.0601 | 2.781 | 0.0623 |
| SEASON | 9 | 7.667 | 0.8519 | 39.434 | < 2e-16 *** |
| DATASET | 2 | 0.003 | 0.0015 | 0.068 | 0.9341 |
| **CLUSTER:SEASON** | 9 | 1.192 | 0.1324 | 6.128 | 1.75e-08 *** |
| **CLUSTER:DATASET** | 2 | 0.021 | 0.0106 | 0.493 | 0.6111 |
| **SEASON:DATASET** | 18 | 0.091 | 0.005 | 0.233 | 0.9997 |
| **CLUSTER:SEASON:DATASET** | 18 | 0.162 | 0.009 | 0.416 | 0.9851 |
| **Residuals** | 1314 | 28.386 | 0.0216 |  |  |
| **PERCENT_H1** (proportion of all influenza A virus detections that were influenza A/H1 viruses) |  |  |  |  |  |
| **CLUSTER** | 2 | 0.48 | 0.242 | 20.221 | 2.24e-09 *** |
| **SEASON** | 9 | 189.66 | 21.074 | 1758.289 | < 2e-16 *** |
| **DATASET** | 2 | 0 | 0 | 0.001 | 0.999 |
| **CLUSTER:SEASON** | 9 | 0.11 | 0.013 | 1.061 | 0.3893 |
| **CLUSTER:DATASET** | 2 | 0.02 | 0.008 | 0.694 | 0.4999 |
| **SEASON:DATASET** | 18 | 0.02 | 0.001 | 0.095 | 1 |
| **CLUSTER:SEASON:DATASET** | 18 | 0.47 | 0.026 | 2.159 | 0.0033 ** |
| **Residuals** | 1310 | 15.7 | 0.012 |  |  |

